# Clinical sequencing of soft tissue and bone sarcomas delineates diverse genomic landscapes and potential therapeutic targets

**DOI:** 10.1101/2021.10.28.21265587

**Authors:** Benjamin A. Nacev, Francisco Sanchez-Vega, Shaleigh A. Smith, Cristina R. Antonescu, Evan Rosenbaum, Hongyu Shi, Cerise Tang, Nicholas D. Socci, Satshil Rana, Rodrigo Gularte-Mérida, Ahmet Zehir, Mrinal M. Gounder, Timothy G. Bowler, Anisha Luthra, Bhumika Jadeja, Azusa Okada, Jonathan A. Strong, Jake Stoller, Jason E. Chan, Ping Chi, Sandra P. D’Angelo, Mark A. Dickson, Ciara M. Kelly, Mary Louise Keohan, Sujana Movva, Katherine Thornton, Paul A. Meyers, Leonard H. Wexler, Emily K. Slotkin, Julia L. Glade Bender, Neerav N. Shukla, Martee L. Hensley, John H. Healey, Michael P. La Quaglia, Kaled M. Alektiar, Aimee M. Crago, Sam S. Yoon, Brian R. Untch, Sarah Chiang, Narasimhan P. Agaram, Meera R. Hameed, Michael F. Berger, David B. Solit, Nikolaus Schultz, Marc Ladanyi, Samuel Singer, William D. Tap

**Author notes:** Equal contribution (Benjamin A. Nacev, Francisco Sanchez-Vega). Co-corresponding authors Corresponding authors: William D. Tap, MD 1275 York Ave. New York, NY 10065, Samuel Singer, MD 1275 York Ave. New York, NY 10065.

## Abstract

The genetic, biologic, and clinical heterogeneity of sarcomas poses a challenge for the identification of therapeutic targets, clinical research, and advancing patient care. Because there are > 100 sarcoma subtypes, in-depth genetic studies have focused on one or a few subtypes. Herein, we report a comparative genetic analyses analysis of 2,138 sarcomas representing 45 pathological entities. This cohort was prospectively analyzed using targeted sequencing to characterize subtype-specific somatic alterations in targetable pathways, rates of whole genome doubling, mutational signatures, and subtype-agnostic genomic clusters. The most common alterations were in cell cycle control and *TP53*, receptor tyrosine kinases/PI3K/RAS, and epigenetic regulators. Subtype-specific associations included *TERT* amplification in intimal sarcoma and SWI/SNF alterations in uterine adenosarcoma. Tumor mutational burden, while low compared to other cancers, varied between and within subtypes. This resource will improve sarcoma models, motivate studies of subtype-specific alterations, and inform investigations of genetic factors and their correlations with treatment response.

## Introduction

Sarcomas are mesenchymal malignancies of the bone or soft tissue that arise in diverse organ sites and display a range of clinical behavior from indolent to aggressive. Sarcomas are also rare tumors, representing < 1% of all malignancies in adults (1). Although the diagnosis and management of sarcomas has slowly improved over the last decade, about 40% of patients with newly diagnosed sarcoma eventually die of disease. One barrier to improving outcomes in sarcoma patients is the cancer’s genomic and biologic complexity, with more than 100 different subtypes now recognized by the World Health Organization (2).

Advances in clinical tumor genomic analyses have improved tumor classification; sarcomas are now classified into two broad genetic groups (3). Sarcomas often have either simple karyotypes, harboring genetic translocations or activating mutations, or highly complex karyotypes, including numerous genomic rearrangements and large chromosomal gains and losses, commonly involving cell cycle genes such as *TP53*, *MDM2*, *RB1*, and *CDK4*. Toward identifying therapeutic targets and designing precision oncology trials based on specific sarcomas’ genetic features, a comprehensive study of soft tissue sarcomas was performed by The Cancer Genome Atlas network, which analyzed 206 samples within 7 common subtypes; rarer ones were represented by as few as 5 cases (4). Analysis of a larger cohort could define the frequency of potentially actionable alterations in rare sarcoma subtypes and broadly compare the frequency of genetic alterations across subtypes to facilitate better diagnostic precision, identify prognostic biomarkers, improve laboratory-based modeling of sarcomas, and generate novel hypotheses on underlying disease mechanisms.

Here, we leveraged an institution-wide tumor genomic profiling initiative to prospectively analyze 2,138 sarcomas encompassing 45 subtypes to identify subtype-specific somatic mutations and copy number alterations and characterize tumor mutation burden (TMB) and microsatellite instability. Paired tumor and normal DNA samples were analyzed using the FDA-cleared Memorial Sloan Kettering-Integrated Mutation Profiling of Actionable Cancer Targets (MSK-IMPACT) next generation sequencing platform (5).

## Results

### Study population characteristics

A total of 2,138 bone and soft tissue sarcoma samples were analyzed. Median patient age was 54 years (range < 1– > 90 years); 1,098 (51.4%) were female. Most were primary tumors; 790 samples (36.9%) were metastases (**Supplementary Table 1**). The analyzed dataset included 45 distinct pathologic entities as assessed by expert sarcoma pathologists. Twenty-two subtypes were represented by ≥ 20 tumor samples and were therefore used as our core subtype set for analyses (**Fig. 1A**). Data from less represented subtypes (**Fig. 1B**) are included in this cohort as a resource. The most common subtypes were gastrointestinal stromal tumor (GIST; n = 395, 18.5%), dedifferentiated liposarcoma (DDLS; n = 167, 5.4%), uterine leiomyosarcoma (ULMS; n = 165, 5.3%), and undifferentiated pleomorphic sarcoma (UPS; n = 145, 4.6%) (**Fig. 1B**). Rare subtypes within the core set include angiosarcoma (ANGS; n = 101, 3.2%), desmoplastic small round cell tumor (DSRCT; n = 53, 1.7%), and perivascular epithelioid cell neoplasms (PECOMA; n = 30, 0.96%) (**Fig. 1B**). As expected, the age distribution varied among subtypes, as did tumor location (**Fig. 1A, Supplementary Table 1**). Among the more common subtypes, myxofibrosarcoma (MFS) had the oldest median age (68 years), whereas embryonal rhabdomyosarcoma (ERMS) had the youngest (8 years). Similarly, sex distribution was not uniform among subtypes (**Fig. 1A**); PECOMA was more common in females (23/30; 76.6%) and DSRCT more common in males (48/53; 90.5%), as was DDLS (males 115/164; 68.8%) (**Fig. 1A**). Among the most common subtypes, survival rate differences were most apparent starting at 3 years post-sequencing. Myxoid/round cell liposarcoma (MRLS) and GIST patients had the highest 3-year survival rates (both > 75%), whereas ANGS and alveolar rhabdomyosarcoma (ARMS) patients had the lowest (34% and 19%, respectively) (**Fig. 1C**).

**Figure 1.**
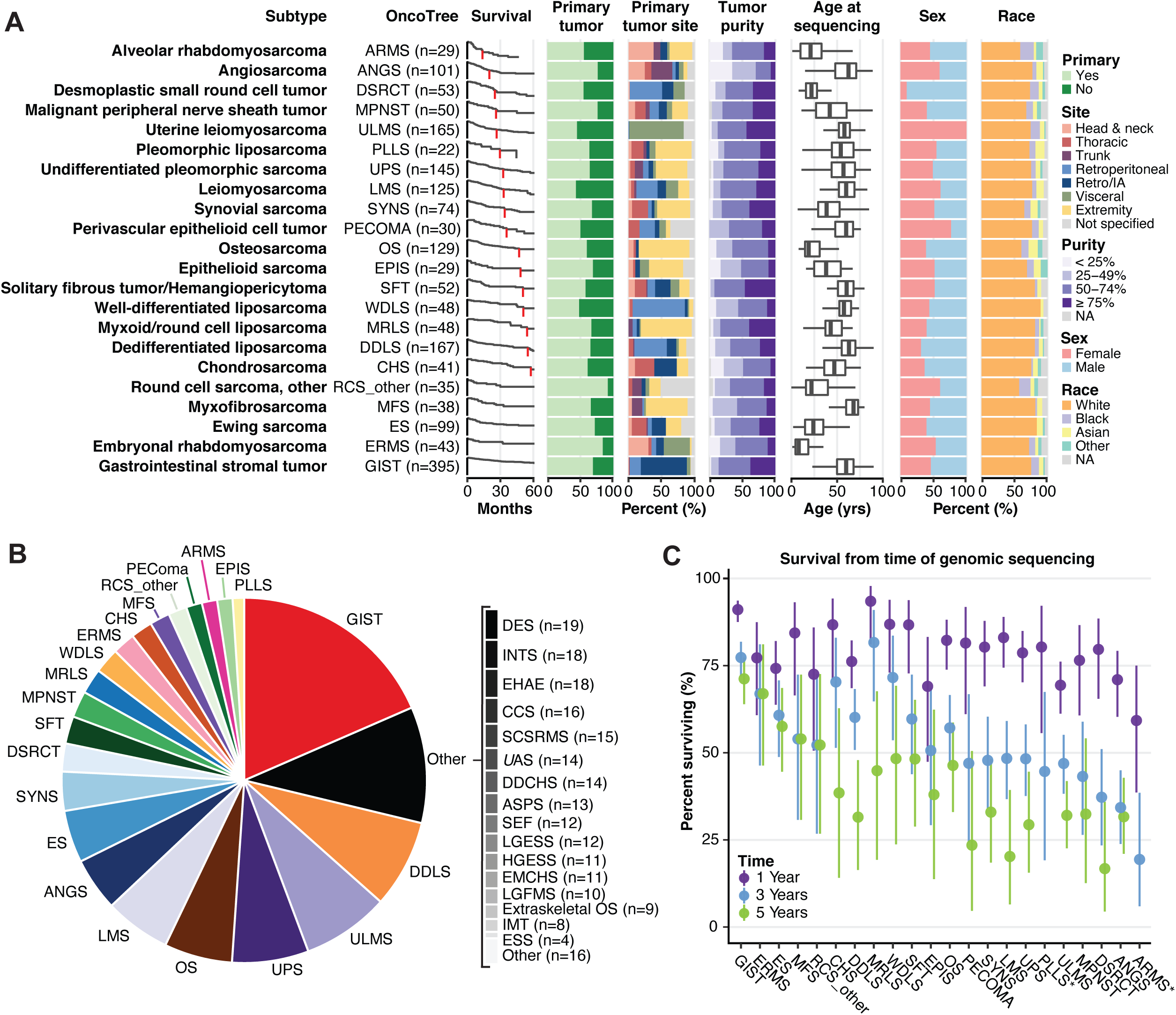
Sample and patient characteristics. This analysis includes 2,138 bone and soft tissue sarcoma samples, each from distinct patients. Subtypes with ≥ 20 samples in the dataset are displayed. **A**, Distribution of number of samples, survival from time of sequencing, sample type (primary or metastatic site), tumor site, sample purity, age, sex, and race in each subtype. **B**, Overall distribution of sample number for the entire cohort. **C**, 1,3, and 5-year survival from the time of sequencing. *, 5-year survival = 0. DES, desmoid tumor; ESS, endometrial stromal sarcoma; INT, intimal sarcoma; LGFMS, low-grade fibromyxoid sarcoma; EMCHS, extraskeletal myxoid chondrosarcoma; HGESS, high-grade endometrial stromal sarcoma; LGESS, low-grade endometrial stromal sarcoma; SEF, sclerosing epithelioid fibrosarcoma; ASPS, alveolar soft part sarcoma; DDCHS, dedifferentiated chondrosarcoma; UAS, uterine adenosarcoma; SCSRMS, spindle cell/sclerosing rhabdomyosarcoma; CCS, clear cell sarcoma; EHAE, epithelioid hemangioendothelioma.

### Subtype-specific mutation analysis

Given the heterogeneity in sarcoma subtypes, their biologic behavior, and clinical presentation, we sought to define the genetics of individual subtypes at both the gene (mutation, gene fusion, and copy number alteration) and functional pathway levels. MSK-IMPACT identified at least one driver mutation in the majority of subtypes (**Fig. 2A, Supplementary Table 2**). Overall, TMB among sarcomas was low, whereas the fraction genome altered (FGA) in most cases was relatively high compared with other cancers, consistent with prior reports (**Fig. 2A**) (4). Both varied greatly among sarcoma subtypes, especially FGA. We performed MutSig and MuSiC analyses to identify significantly recurrently mutated genes in each subtype (**Fig. 2B**). As expected, *TP53* and *RB1* were significantly altered across multiple subtypes, but at markedly different frequencies. Within GIST, we identified several frequently mutated genes in addition to previously known drivers such as *KIT*, *SDHA*, and *PDGFRA* (6). These included the histone methyltransferase *SETD2* (4%), the MYC binding partner and transcription factor *MAX* (4%), and *MGA* (3%), which binds the MAX-MYC complex (7).

**Figure 2.**
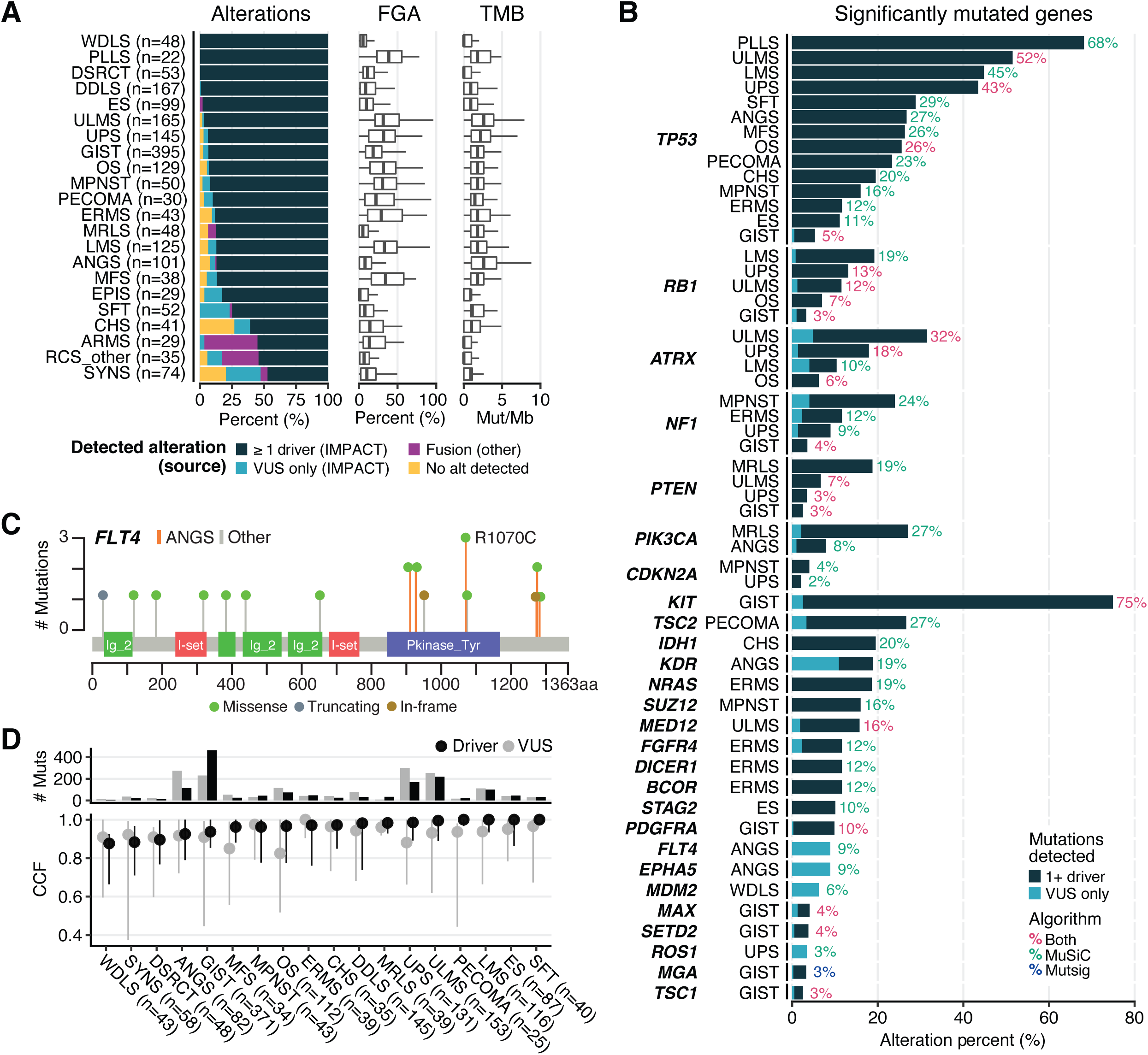
Mutation analysis by subtype. **A**, Alteration type and frequency, fraction of the genome altered (FGA) and tumor mutation burden (TMB) by subtype. Oncogenic fusions detected by MSK-IMPACT are classified as drivers. **B**, Significant mutations were identified in all subtypes with n ≥ 20 in our dataset using both MutSig and MuSiC analysis. Percentages indicate the percentage of samples with an oncogenic mutation in the corresponding gene. **C**, *FLT4* mutation type, frequency, and location in ANGS vs. other subtypes. **D**, Cancer cell fraction (CCF) and number of mutations for driver mutations and VUS by subtype.

Additional subtypes with recurrently mutated genes of potential biologic or clinical relevance included ANGS (n = 101), in which we identified recurrent mutations in receptor tyrosine kinases involved in angiogenesis including *KDR* (VEGFR2; 19%) and *FLT4* (VEGFR3; 9%) as well as another receptor tyrosine kinase, *EPHA5* (9% of cases). The mutations in *EPHA5* and *FLT4* were all variants of unknown significance (VUS). The VUS in *FLT4* all affect the kinase domain or the C-terminus, implying a possible functional consequence (**Fig. 2C**). In Ewing sarcoma (ES; n = 99), 10% of samples carried mutations in the cohesion complex component *STAG2*, confirming prior reports (8). In ULMS, *MED12*, a member of the transcription elongation complex, was altered in 16% of cases, most frequently missense mutations at glycine 44, as reported previously (**Fig. 2B**) (9).

In PECOMA, SFT, LMS, ULMS, and ES, driver mutations were represented in a cancer cell fraction (CCF) of close to 1.0, suggesting that these represent a large clonal population (**Fig. 2D**). The CCF for VUS was overall similar to that of drivers within most subtypes, with the exception of MFS, OS, and UPS, which suggests that in some cases these VUS could have an unrecognized function, calling for further studies to determine their roles in oncogenesis and progression.

### Copy Number Alterations by Subtype

As many sarcomas are driven by copy number alterations, we analyzed these changes across the whole cohort, including in subtypes not classically thought to be driven by them (**Fig. 3**). For instance, in GIST patients (evaluable n = 371), there were frequent copy number loss events involving chromosomes 1, 14, 15, and 22 **(Fig. 3A**). Translocation-driven sarcomas, e.g. ES, DSRCT, and SYNS, exhibited highly recurrent copy number changes, indicating that there may be additional relevant genetic events beyond the driver translocations **(Fig. 3B**). We identified a diversity of chromosome arms (e.g. 5p, 8q, and 10p) that were recurrently affected by copy number variation across multiple common subtypes (**Fig. 3B**). Of note, 12q amplifications in DDLS and WDLS patients were not wide enough to be called in arm-level analysis (**Fig. 3B**, left), though they were clearly observed as a strong focal event in copy number profiles (**Fig. 3A, B**, right). In most cases, these arm-level copy number events were not linked to a specific gene. However, there were some exceptions including significant gains of *MYC* on chromosome 8q24 in OS, EPIS, ERMS, and ANGS, as well as significant gain of a negative regulator of NF-kB signaling, *TNFAIP3*, in DDLS (**Fig. 3B**). As expected, we observed more widespread copy number changes in classically copy number-driven subtypes such as LMS, ULMS, MFS, and OS compared with the rest of the cohort.

**Figure 3.**
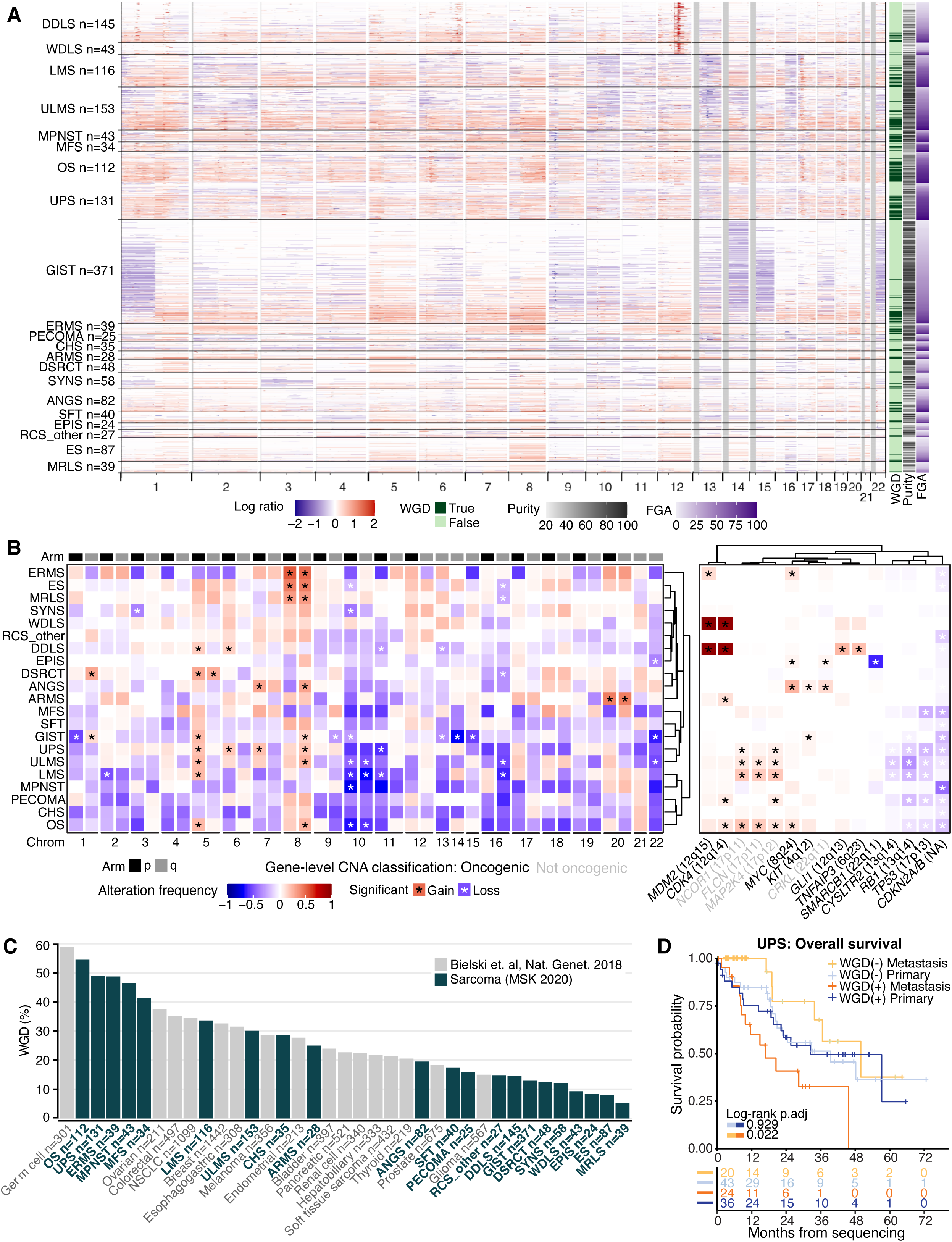
Copy number changes by subtype. **A**, Individual sample copy number variation (CNV) across the genome for each subtype. Whole genome doubling (WGD), fraction genome altered (FGA), and purity are shown at right. **B**, Aggregate arm-level (left) and gene-level events (right) grouped by subtype. *, significant change based on Bonferroni corrected p-values. Oncogenic vs. non-oncogenic CNV classifications according to OncoKB. **C**, Frequency of WGD by subtype (green) compared to other cancers with ≥ 200 samples available for comparison (gray) (12). **D**, Overall survival based on WGD status within primary and metastatic samples.

Despite sharing *CDK4* and *MDM2* amplification events, DDLS is more aggressive than WDLS and has increased risk for distant spread (10). Therefore, we compared rates of amplifications between WDLS and DDLS, and found greater rates of amplification of the oncogenes *GLI1* (8.5% vs. 25.3%), *TERT* (6.3% vs. 14.4%), and *JUN* (0% vs. 13.8%) in DDLS. The Jun transcription factor positively regulates the expression of cyclin D1, a CDK4/6 cyclin partner, and amplification of JUN in DDLS is associated with a more aggressive phenotype (4), calling for investigation of whether *CDK4* and *JUN* co-amplification drives progression to DDLS or modulates response to *CDK4* inhibition. Amplification of the GLI1 transcription factor, downstream of Sonic Hedgehog (Shh) signaling, has previously been reported (11); this confirmation furthers rationale for studying Shh pathway inhibition in DDLS. *GLI1* amplification and *JUN* amplification were mutually exclusive.

OS, UPS, ERMS, and MPNST had high frequencies of WGD, all around 50%, ranking among the highest even among a wide variety of cancers for which WGD was previously analyzed (**Fig. 3C; Supplementary Fig. S1A**) (12). In keeping with the notion that MFS is on a genetic continuum with UPS (4), UPS and MFS had similar WGD frequencies. Despite being copy number variation (CNV)-driven, WDLS and DDLS had lower rates of WGD frequency, as did many translocation-driven subtypes including SYNS, ES, DSRCT, and MRLS. In sarcomas, WGD was associated with worse overall survival among metastatic (p = 0.042) but not primary cases (p = 0.391; **Supplementary Fig. S1B)**. Among specific subtypes, WGD was associated with worse overall survival (from time of sequencing) in metastatic UPS (p = 0.022; **Fig. 3D**), but not MFS (p = 0.78; **Supplementary Fig. S1C**).

### Commonly Disrupted Pathways in Sarcoma

Pathway-specific analyses within each sarcoma subtype for which ≥ 20 samples were available (**Fig. 4A**, genes in each pathway listed in **Supplementary Table 3)** revealed that a number of pathways important in carcinomas were infrequently altered in sarcoma, including TGFβ, WNT, Hippo, Notch, and NRF2 (**Fig. 4A**, right panel). By contrast, the cell cycle and *TP53* pathways were altered in at least half of samples in 8 of the 22 most common subtypes. For instance, DDLS and WDLS demonstrated cell cycle or *TP53* pathway alterations in 214/215 (99%) of samples, most commonly through co-amplification of *CDK4* and the E3 ubiquitin ligase that targets p53 for degradation, *MDM2* (**Fig. 4A**) (13, 14). Many of the sarcomas with infrequent alterations (< 10%) in the cell cycle and *TP53* pathways were driven by translocations (e.g. MRLS, DSRCT, SYNS) or alteration in the SWI/SNF remodeling complex (epithelioid sarcoma [EPIS]) (**Fig. 4A**), highlighting a distinct mechanism of pathogenesis. An exception was solitary fibrous tumor (SFT), which is driven by the *NAB2-STAT6* fusion oncogene, and has oncogenic *TP53* alteration in 28% of cases (**Fig. 4A**) (15).

**Figure 4.**
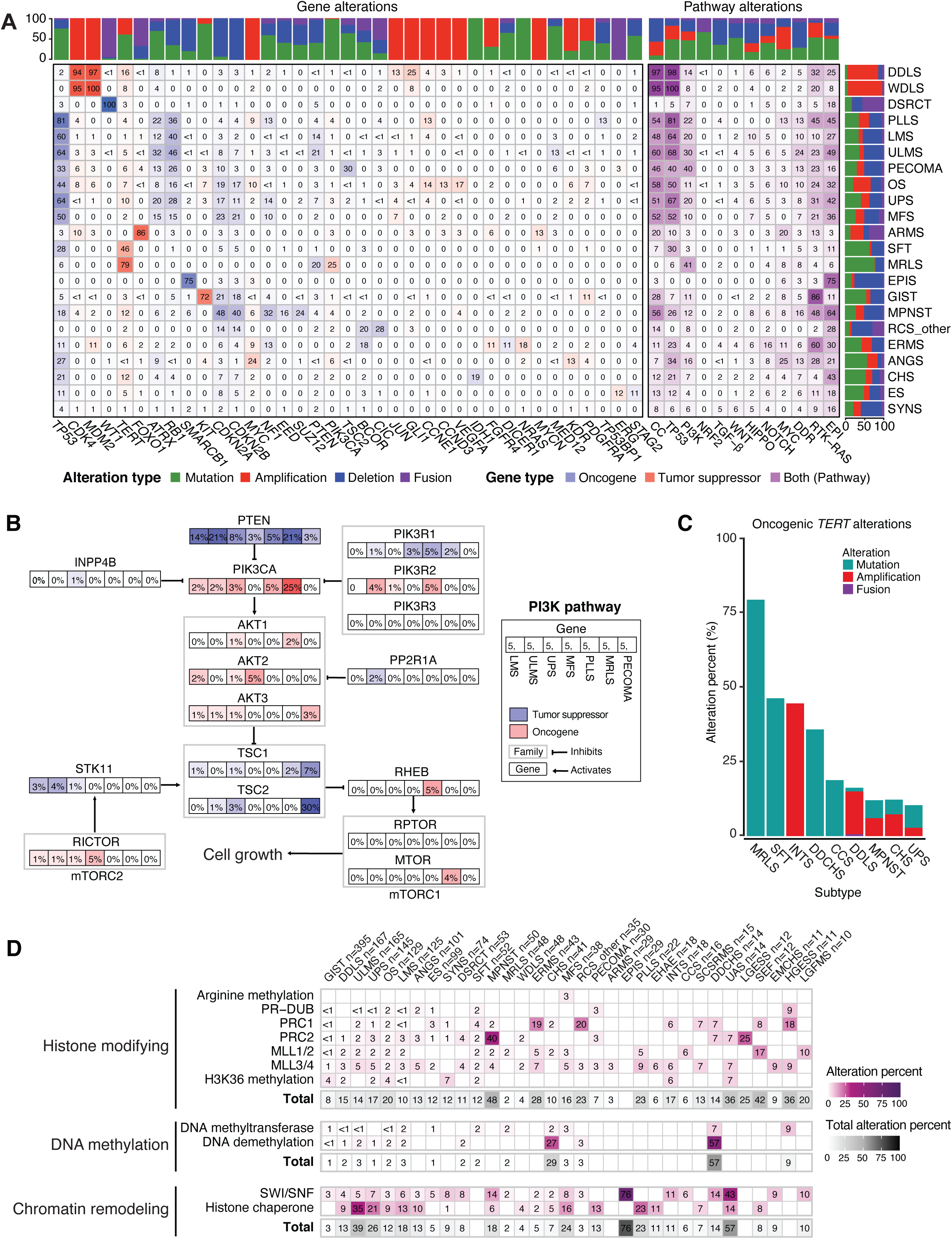
Integrated pathway analysis. **A**, Oncogenic alterations within each of 12 pathways with relevance to cancer biology in each subtype. Numbers in each cell indicate percentage of samples harboring alterations. Stacked bar graphs indicate the distribution of the type of oncogenic alteration per gene or pathway (top) or subtype (right). CC, cell cycle; EPI, epigenetic. **B**, PI3K pathway alterations in specific subtypes. The percentage of samples with an alteration in a specific gene in each subtype is indicated in each box. **C**, Oncogenic *TERT* promoter alterations in each of the 9 most altered subtypes. **D**, Oncogenic epigenetic pathway alterations by subtype, grouped by complex and/or biochemical function of the encoded protein. Totals include all alterations in genes that belong to a parent category, not only those affecting specific complexes listed.

The PI3K pathway was frequently altered in some of the most prevalent subtypes in our dataset including MRLS (41%), PECOMA (40%), ULMS (30%), pleomorphic liposarcoma (PLLS; 22%), UPS (20%), and soft tissue leiomyosarcoma (LMS; 20%) (**Fig. 4A**, right panel). Among these subtypes, *PTEN* and *PIK3CA* were the most frequently affected genes except in PECOMA where *TSC2* loss of function alterations were most common (30%) (**Fig. 4A, 4B**). *PTEN* loss of function alterations predominate in LMS and ULMS (14% and 21%, respectively), whereas in MRLS, *PIK3CA* mutations were most frequent, occurring in 25% of cases, consistent with our prior findings (16). In MRLS, *PTEN* loss is observed in 21% of cases, some of which were concurrent with *PIK3CA* mutations (4 *PIK3CA* mutations in 10 *PTEN* loss cases). In contrast, in UPS, *PTEN* alterations were identified in 8% of samples and *PIK3CA* in 3%; only 1 of the 11 cases with a *PTEN* alteration had a concurrent *PIKC3A* mutation. Notably, *PTEN* loss of function has also been proposed as a predictor of non-response to immune checkpoint inhibition in ULMS (17).

Because a pan-cancer MSK-IMPACT analysis identified *TERT* promoter mutations in a subset of sarcomas (18), we investigated their frequency as a function of sarcoma subtype (**Fig. 4C**). We identified oncogenic *TERT* amplifications in 44% (8/18) of intimal sarcoma (INTS) and *TERT* promoter mutations in 79% (38/48) of MRLS, 46% (24/52) of SFT, and 35% (5/14) of dedifferentiated chondrosarcoma (DDCHS). In DDLS, oncogenic *TERT* promoter alterations were present in 16% of samples (27/167) and were almost entirely amplifications (n = 24). *TERT* copy number alterations have not yet been described in INTS, perhaps due to the low incidence of this rare subtype. The *TERT* locus is distinct from that of the *MDM2* and *CDK4* amplifications (19) that are hallmarks of INTS, implicating *TERT* amplification as a potential independent contributor to pathogenesis.

Alterations in DNA damage repair (DDR) pathway genes have been associated with development of sarcomas (20), and are of particular clinical interest as PARP inhibition has activity in select carcinomas with homologous recombination deficiency and immune checkpoint blockade has activity in certain tumors with microsatellite instability (21, 22). Our analysis of DDR pathway alterations found that 9.6% of all samples harbored an oncogenic somatic alteration in a DDR pathway. Among subtypes with more than 20 samples, the frequency of DDR gene alterations was highest in ULMS (24%), MPNST (16%), PLLS (13%), PECOMA (13%), ANGS (13%), LMS (10%), and OS (10%) (**Fig. 4A**, right panel). The most frequently altered genes across subtypes were *BRCA2* (1.4% of all samples)*, RAD51B* (1.1%)*, CHEK2* (1.0%) *ATM* (0.9%)*, FANCA* (0.6%), and *RAD51* (0.6%). Consistent with a previous report in uterine sarcomas (23), nearly half of *BRCA2* (41%) and *RAD51B* (47%) alterations occurred in sarcomas of uterine origin, with *RAD51B* or *BRCA2* each mutated in 7% of ULMS cases. Similarly, 35% of the 14 uterine adenosarcomas also had an altered DDR gene, all deep deletions. Five percent of ANGS had oncogenic mutations and another 5% had a VUS in *ATM*. Given the association of ANGS with prior ionizing radiation, *ATM* mutations may represent a convergent pathogenic mechanism for accumulation of DNA damage. Of the 15 sarcomas (0.7%) with an altered mismatch repair (MMR) gene (*MLH1, MSH2, MSH6,* or *PMS2*), one (LMS) was microsatellite instability (MSI)-high by MSISensor and had a high TMB.

Epigenetic dysregulation contributes to the pathogenesis of several sarcoma subtypes (24). In SYNS, EPIS, malignant rhabdoid tumors, and MPNST, this occurs through alterations of chromatin-remodeling and -modifying complexes; in chondroblastoma, CHS, UPS, giant cell tumors of bone, and osteosarcoma through oncogenic histone mutations. In light of emerging pharmacologic strategies to study and therapeutically target epigenetic regulatory proteins, we identified sarcoma subtypes characterized by epigenetic pathway alterations (**Fig. 4A**; **Supplementary Table 3**) (25). As expected, 75% of EPIS had loss-of-function deletions, truncating mutations, or intragenic fusions in *SMARCB1*. In addition, the epigenetic pathway was one of the most altered pathways among the highly prevalent subtypes in our dataset. Pathogenic alterations in epigenetic pathway genes (**Supplementary Table 3**) were observed in 64% of MPNST, 49% of ULMS, 45% of PLLS, 43% of CHS, 42% of UPS, 36% of MFS, and 32% of OS (**Fig. 4A**). By contrast, these alterations were infrequently observed (< 10%) in WDLS, ARMS, and MRLS, suggesting subtype specificity.

We determined the association with specific subtypes of epigenetic pathway genes contributing to a specific biochemical function (e.g. DNA methylation, chromatin remodeling) and complex (e.g. PRC1, PRC2, MLL3/4) (**Fig. 4D**; **Supplementary Fig. 2; Supplementary Table 4**). Genes involved in histone modification were altered in 48% of MPNST, 42% of sclerosing epithelioid fibrosarcoma (SEF), 36% of uterine adenosarcoma (UAS), and 36% of high-grade endometrial stromal sarcoma (HGESS). ERMS had frequent alterations in in the transcriptional co-repressor and non-canonical PRC1 complex member *BCOR* (19% total, 16% oncogenic), which were mutually exclusive with *DICER1* alterations (12% oncogenic). Both alterations were more prevalent in our population than in prior studies (26, 27).

Genes involved in chromatin remodeling were altered at similarly high frequencies: 76% of epithelial sarcoma (EPIS), 39% of ULMS, 26% of UPS, 24% of MFS, and 18% of MPNST. In a significant portion of these cases, the histone chaperone *ATRX* drove these high rates (**Supplementary Fig. 2**). We also note the unexpected finding that UAS (n = 14), a rare sarcoma subtype, had oncogenic alterations in genes encoding subunits of the SWI/SNF chromatin remodeling complex in 43% of patients, with *ARID1A* and *PBRM1* most frequently affected (**Fig. 4D**, **Supplementary Fig. 2**). Interestingly, UAS also had alterations in histone-modifying genes in 36% of cases.

As epigenetic alterations are more frequent in DDLS (25%) than WDLS (8%) and we have previously found epigenetic dysregulation to contribute to DDLS (28), we further examined differences between DDLS and WDLS. Histone-modifying and histone chaperone/chromatin-remodeling alterations occur in 15% and 13% of DDLS cases, respectively, compared with 4% each of WDLS (**Fig. 4D**). This suggests that loss of epigenetic regulation could be an important contributor to dedifferentiation.

We also examined epigenetic pathways without filtering for alterations already established as oncogenic, which is a strategy we recently employed to generate hypotheses in an analysis of genetic alterations in OS (**Supplementary Fig. 3**) (29). This analysis identified the histone methyltransferase *KMT2D/MLL4* as more frequently altered in MFS (16%) compared with other subtypes including the closely related UPS (6%) (4). Histone-modifying enzymes were altered in 20% of SYNS, among which *KMT2B* and *SETD2* were altered in 6% and 7% of samples, respectively. We also found that the transcriptional corepressor *NCOR1*, which complexes with *HDAC3* and other deacetylases to regulate the activity of transcription factors such as the retinoic acid receptor and thyroid hormone receptor (30), was altered in 10% of ULMS, 19% of LMS, and 21% of OS, mostly through amplification. *NCOR1* has also been shown to regulate transcription factors important in mesenchymal lineages including the MEF2 family and PPARγ, which regulate myo- and adipogenesis, respectively (31). Several other genes within the same cytoband as *NCOR1*, 17p12-p11.2, were co-amplified, including *FLCN*, *MAP2K4*, *AURKB*, and *ALOX12B* (**Supplementary Fig. 4A**). Amplifications of *MYOCD*, whose genomic location is within a region previously found to be amplified in LMS (32), were not detected because this gene is not represented on the MSK-IMPACT panel. Except for *ALOX12B*, the 17p copy number gains of MSK-IMPACT-assessed genes were associated with increased gene expression in the sarcoma TCGA analysis (**Supplementary Fig. 4B**). Thus, one or more of these genes could play a pathogenic role.

### Mutual Exclusivity and Co-occurrence

To better understand how gene- and pathway-level alterations interact, we analyzed their co-occurrence and mutual exclusivity (**Fig**. **5A**). As expected, *KIT* and *PDGFRA* alterations were mutually exclusive in GIST and *CDK4* and *MDM2* co-occurred in DDLS. In OS, *KDR* alterations co-occurred with *KIT* and *PDGFRA,* as did the latter two with each other, suggesting dysregulation of signaling through these 3 RTK genes located at the 4q12 locus (29). *TP53* alterations were mutually exclusive with *CDKN2A/B* in GIST and ULMS, but not in UPS. In SFT and ES, *TP53* alterations co-occurred with *STAG2* and *TERT* alterations, respectively, suggesting context dependence for alterations co-occurring with *TP53*. In UPS, *ATRX* and *NF1* alterations, which are mostly loss-of-function events, were mutually exclusive, suggesting biologically different subgroups.

**Figure 5.**
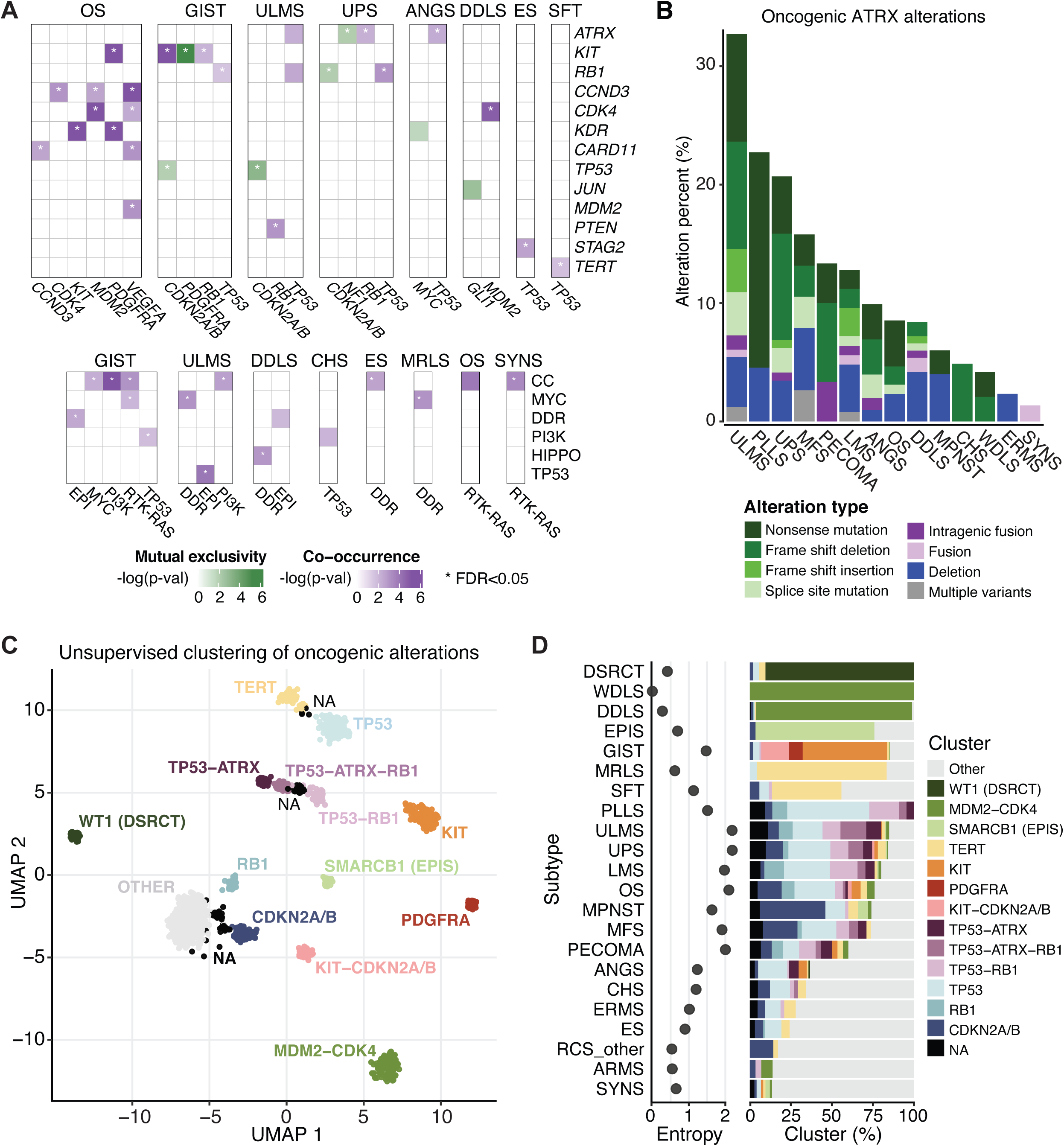
Mutual exclusivity, co-occurrence, *ATRX* alterations, and unsupervised clustering based on genetic signatures. **A**, Co-occurrence and mutual exclusivity of gene- (top) and pathway-level (bottom) alterations in each subtype. **B**, Frequency and types of oncogenic *ATRX* alterations in each of the 14 most altered subtypes. **C**, Unsupervised clustering of samples based on oncogenic alteration patterns. D, Subtype-specific cluster associations and entropy scores.

At the pathway level, cell cycle and DDR pathway alterations significantly co-occurred (false discovery rate [FDR] < 0.05) with those in other pathways. For instance, cell cycle alterations co-occurred with MYC pathway alterations in GIST, with PI3K pathway alterations in GIST and ULMS, and with RTK/RAS alterations in GIST, OS, and SYNS (**Fig. 5A**). DDR pathway alterations co-occurred with MYC pathway alterations in ULMS and MRLS, epigenetic pathway alterations in GIST and DDLS, Hippo pathway alterations in DDLS, and cell cycle alterations in ES. There were no examples of significant mutual exclusivity at the pathway level.

### *ATRX* Alterations across Subtypes

*ATRX* stood out across subtypes as frequently affected by loss-of-function events (**Fig. 5B**); this gene was altered in ≥ 10% of cases in 7 subtypes: ULMS, PLLS, UPS, MFS, PECOMA, LMS, and ANGS. In ULMS, which had the highest rate of *ATRX* alterations, the frequency was roughly 1 in 3 cases. That *ATRX* loss-of-function events occur in both copy number- and translocation-driven subtypes, although at lower frequency in the latter, raises the possibility that they may serve a fundamental role in the biology of a molecular subset of these subtypes. *ATRX* loss-of-function mutations were more frequent than deletion events, independent of subtype, despite the overall low mutation rate. Our analysis also captured intra- and intergenic *ATRX* fusion events.

### Unsupervised Clustering of Subtypes

To assess genetic similarities among subtypes, we grouped samples on the basis of genetic alterations by unsupervised clustering (**Fig. 5C**), which generated 15 distinct clusters subsequently named according to their prevailing subtype and/or genetic feature. Some subtypes and clusters were closely associated (**Fig. 5D**). These included EPIS and the *SMARCB1* cluster, DSRCT and *WT1,* and WDLS and DDLS with *MDM2*-*CDK4.* These groupings largely reflect known or presumed drivers in these subtypes and reinforces their central roles therein.

Other clusters were heterogeneous and comprised of many subtypes. One such cluster was associated with frequent alterations in *TERT*, which dominated by MRLS and SFT, but also included small populations of other sarcoma subtypes. Another cluster lacked any predominantly altered gene. This ‘other’ cluster included the majority of samples in some histotypes (e.g. ANGS and CHS) but also included samples from multiple subtypes that are represented more commonly in other clusters (e.g. LMS and UPS), suggesting that they may be genetic outliers among those subtypes.

Notably, alterations in *TP53*, thought of as a canonical driver in many sarcomas, were associated with 4 closely related clusters, each with a distinct association with co-occurring *ATRX* and/or *RB1* alterations. In contrast, the *RB1-*altered/*TP53*-WT (“*RB1*”) group clustered distantly from the *TP53*-altered groups and was more closely related with *CDKN2A*/*B* and ‘other’ groups. ULMS, PLLS, and UPS were represented in both the *TP53-*altered groups and the *RB1-*altered/*TP53*-WT group.

For each subtype we also assigned an entropy score with respect to the clustering assignments (**Fig. 5D**). WDLS, DDLS, and DSRCT had the lowest entropy, suggesting relatively uniform genomic profiles within each subtype, whereas ULMS, UPS, and OS had high entropy, suggesting that these pathologically defined entities harbor multiple distinct genetic variants.

### Tumor Mutational Burden, Microsatellite Instability, and Mutational Signatures

Two recent immune checkpoint blockade trials in sarcoma demonstrated low overall response rates, though rates varied among subtypes (33, 34). Thus, predictive biomarkers for response to checkpoint blockade are needed to deconvolute this heterogeneity and aid in the design of future clinical trials. As microsatellite instability predicts response to pembrolizumab (21), and tumors with high TMB are more likely to respond to immune checkpoint blockade (35), we determined MSI status and TMB for each subtype (**Fig. 6A and 6B, Supplementary Table 1**).

**Figure 6.**
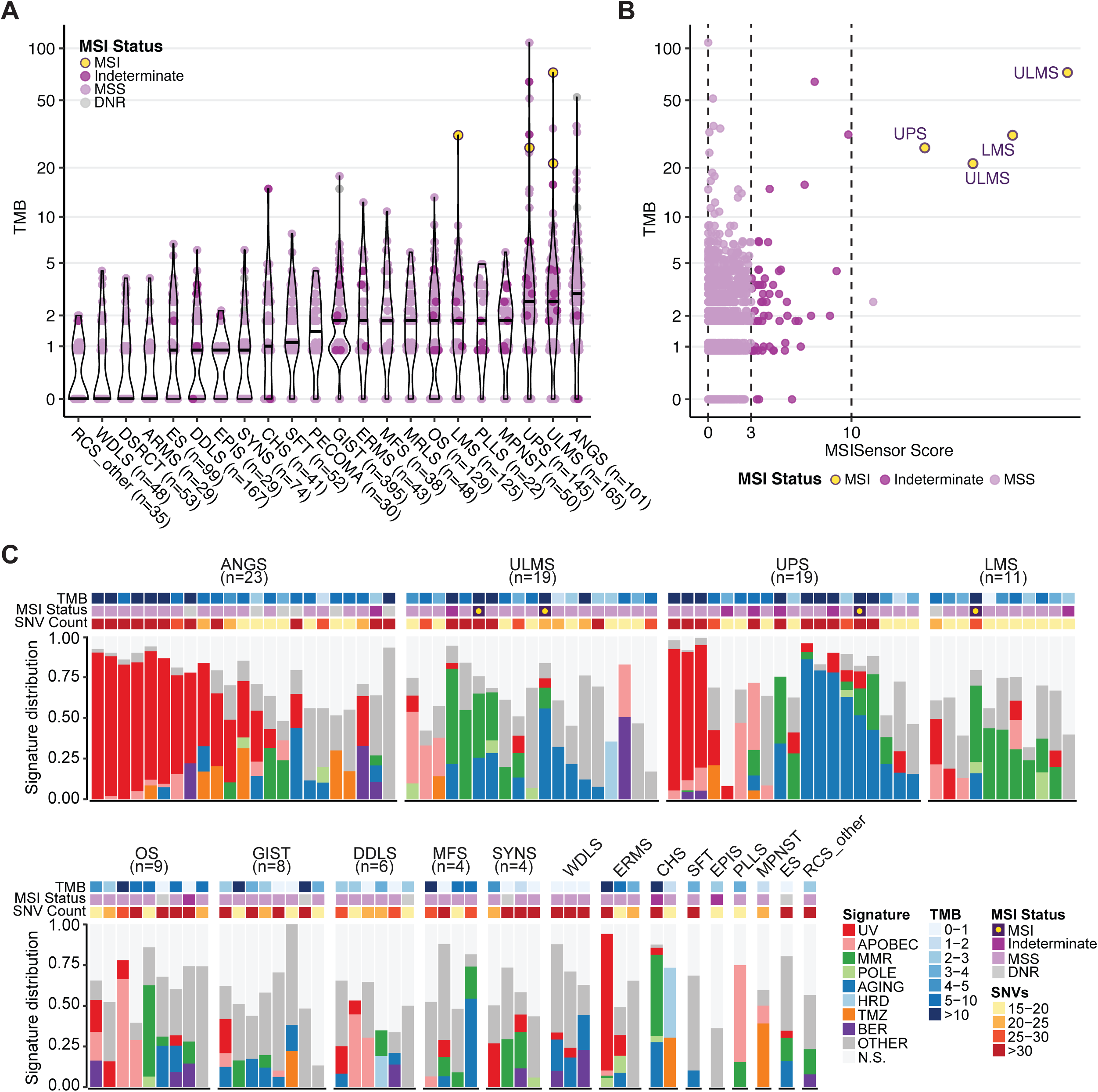
Tumor mutation burden (TMB), microsatellite instability (MSI) status, and mutational signatures. **A**, Distribution of TMB (mutations/Mb) by subtype with sample-level MSI status. **B**, MSISensor score versus TMB for the cohort with sample-level MSI status. **C**, Mutational signature profile of each evaluable sample. Each bar represents the signature distribution of an individual sample, grouped by subtype. TMB, MSI status, and single nucleotide polymorphism (SNV) count are shown above.

While the median TMB for sarcomas was low compared to many carcinomas (4), there is considerable heterogeneity within and between the more common subtypes in our cohort (inter-subtype median range 0.9–3.0) (**Fig. 6A**). The median TMB was greatest in ANGS (3.0), UPS (2.6), and ULMS (2.6) and lowest in WDLS (0.9), EPIS (0.9), and RCS (other) (0.9). However, in certain subtypes, the distribution of TMB had a long upper tail and was skewed towards higher TMB (**Fig. 6A**). TMB was ≥ 5 mut/Mb in 25% of ANGS, 15% of ULMS and UPS, and 13% of ERMS. Only two subtypes had ≥ 5% of samples with a TMB of ≥ 10 mut/Mb: ANGS (7.6%) and UPS (6.7%).

Only 5 of 1893 samples evaluable for MSI status were MSI-high (by MSIsensor score ≥ 10), including one UPS, one LMS, and 3 ULMS (**Fig. 6B**). Of these, 4 were confirmed to be MSI-high by a conventional PCR-based MSI assay. MSIsensor scores varied widely between subtypes (**Supplementary Table 1**). Overall, while microsatellite instability corresponded with high TMB, the inverse was not true.

To understand mechanisms contributing to extensive genetic alterations, we examined mutational signatures in samples with ≥ 15 single nucleotide variants (SNVs) (**Fig. 6C**). A UV mutational signature was observed in a subset of ANGS and was most strongly observed in samples at the highest end of the TMB spectrum within that group. Of these 16 samples, 11 had a head and neck primary site. Interestingly, a subset of UPS also harbored a UV signature and a higher TMB, while another subset of highly mutated UPS was dominated by an aging signature, suggesting alternative mechanisms for high TMB within UPS.

### Subtype-specific actionable alterations

Toward improved detection of targetable alterations for each subtype and patient, we analyzed genetic alterations by actionability according to OncoKB (**Fig. 7**) (36). As expected, level 1 alterations, defined as FDA-recognized biomarkers for response to an FDA-approved drug, were most frequent in GIST due to *KIT* and *PDGFRA* mutations. Similarly, level 1 *SMARCB1* deletion was noted in 66% of EPIS. Level 2 alterations, defined as guideline-supported standard-of-care biomarkers for an FDA-approved drug, were seen in > 90% of WDLS and DDLS related to *CDK4* amplification. In the same subtypes, *MDM2* amplifications in > 90% of cases were deemed Level 3A, for which compelling evidence supports use as a predictive biomarker for an existing drug. Many other observed alterations were classified as Level 3B, defined as standard-of-care or investigational biomarkers that predict response to an FDA-approved or investigational drug in another cancer. Notable examples included a combined 37% prevalence of actionable *TSC1/2* deletions in PECOMA, *IDH1/2* alterations in 27% of CHS, and targetable PI3K pathway (*PIK3CA*, *ATK1*, *MTOR*, or *TSC1*) alterations in a collective 31% of MRLS. Notably, 21% of MRLS cases had Level 4 *PTEN* deletions, for which compelling biological evidence supports their use as a predictive biomarker. Other intriguing Level 4 alterations included somatic *NF1* deletions in MPNST (32%), UPS (14%), ERMS (14%), and PLLS (14%), and *CDKN2A* deletions in many subtypes at a rate of up to 48% as seen in MPNST.

**Figure 7.**
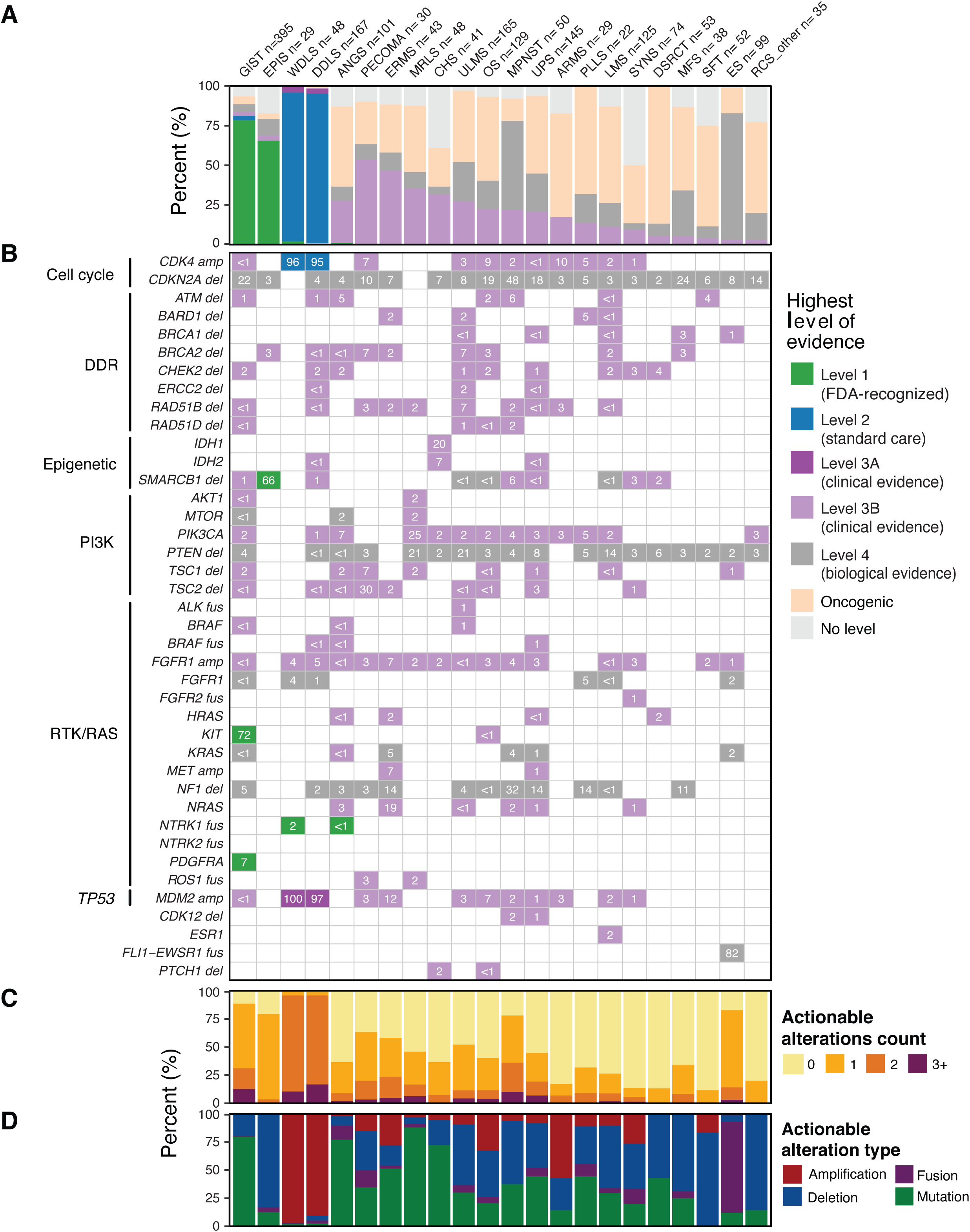
Actionability of mutations by subtype and gene. For each of the 22 most common subtypes: **A**, Frequency of actionable alterations by level of evidence. **B**, Actionable alterations in individual genes, grouped by pathway. Numbers in each cell represent the percentage of samples with actionable alterations in that gene. **C**, Number of actionable alterations per sample. **D**, Frequency of actionable alterations classified by alteration type.

## Discussion

To better understand genetic heterogeneity in sarcomas, we analyzed prospectively generated tumor next generation sequencing data from a cohort of 2,138 sarcoma samples representing 45 histological subtypes. Across all subtypes, the most common alterations we identified were in cell cycle control and *TP53*, receptor tyrosine kinases/PI3K/RAS, and epigenetic regulators. Previously unreported subtype-specific associations included *TERT* amplification in intimal sarcoma and SWI/SNF complex alterations in uterine adenosarcoma. Tumor mutation burden varied widely between and within subtypes.

Epigenetic pathway mutations frequently occurred in many subtypes in our cohort, in keeping with an emerging recognition of epigenetic dysregulation as an important factor in the pathogenesis of sarcomas (24). A common epigenetic pathway alteration was amplification of *NCOR1*, particularly in ULMS, LMS, and OS. *NCOR1* is a transcriptional corepressor that regulates transcription factors specific to mesenchymal lineages and can suppress differentiation when overexpressed (31, 37). If amplification of *NCOR1* correlates with increased protein levels in these sarcomas, which RNA sequencing data suggests it may, this could lead to altered differentiation and transcriptional programs. Moreover, since the activity of *NCOR1* is modulated by PI3K/Akt-mediated control of nuclear localization, both inhibition of that pathway and of *HDAC3* warrant further exploration as potential therapeutic strategies in *NCOR1*- amplified ULMS, LMS, or OS (38).

In uterine adenosarcoma, we identified genetic alterations in the SWI/SNF chromatin remodeling complex in 43% of cases, mostly loss-of-function alterations in *ARID1A* or *PBRM1*. Uterine adenosarcoma is a rare subtype composed of both sarcomatous stroma and benign epithelium, which can behave aggressively, especially in the setting of sarcomatous overgrowth (39). Given the role of epigenetic regulation in determining differentiation, impaired SWI/SNF function could contribute to this phenotype. Histone mutations have been observed in ovarian carcinosarcoma, suggesting that epigenetic dysregulation may be a common mechanism for impaired lineage commitment in Müllerian tumors (40). Given synthetic lethality between the loss of the SWI/SNF component *SMARCB1* in epithelioid sarcoma and EZH2 inhibition with the now FDA-approved drug tazemetostat, EZH2 inhibition may represent a future therapeutic strategy in uterine adenosarcoma (41).

We also analyzed genes involved in maintenance of telomeres, whose tumor-suppressive function is dependent on the silencing of *TERT*, a reverse transcriptase and core component of telomerase. Mutations in the *TERT* promoter, first identified in melanoma, lead to increased transcription of the TERT gene (42, 43). Within our cohort, *TERT* amplification occurs in 44% of intimal sarcomas, which to our knowledge has not been previously reported. Whether this amplification leads to increased expression of the TERT gene product should be investigated, as *TERT* overexpression is known to be oncogenic in certain contexts (44). In addition, our data validate prior findings of *TERT* mutations in MLPS and SFT. However, the rate in SFT was greater than observed in prior studies (45), which may be explained by differences in disease aggressiveness, as *TERT* mutations associate with worse prognosis (46).

While we included *ATRX* in the epigenetic pathway gene list owing to its fundamental role, along with *DAXX*, as a histone variant H3.3 chaperone, *ATRX* also participates in other pathways including telomerase-independent alternative lengthening of telomeres (ALT), which has been observed in a number of soft tissue sarcomas including UPS and liposarcoma (47). Because UPS and liposarcoma also harbor *TERT* alterations in a largely non-overlapping pattern, these sarcomas may acquire the ability to aberrantly maintain telomeres through multiple independent mechanisms. In addition to epigenetic and ALT functions, *ATRX* helps maintain genomic integrity (48). Because of the diversity of the physiologic functions of *ATRX,* the role(s) of *ATRX* alterations in sarcomagenesis are difficult to predict *a priori*. Thus, developing tools such as patient-derived cell lines and xenografts to study the impact of these alterations on *ATRX*-dependent functions will be informative. Given the relative frequency of *ATRX* alterations and the inclusion of *ATRX* on MSK-IMPACT and other tumor sequencing platforms, such investigations are eminently feasible.

Toward identifying predictive biomarkers for response to immune checkpoint blockade in sarcoma, we analyzed the distribution of MSI-H and high TMB, which are associated with response to these agents in other solid tumors. Almost none of the samples had microsatellite instability and there were relatively few samples with high TMB. However, the upper tail of TMB was relatively long in certain subtypes such as UPS, ANGS, and ULMS. Moreover, we do not yet know whether the TMB cutoff of 10 mutations per megabase, which defines high TMB for carcinomas and predicts response to immune checkpoint blockade, is the appropriate threshold for TMB as a predictive biomarker in mesenchymal neoplasms, let alone specific sarcoma subtypes. Indeed, recent work suggests that the highest quintile of TMB within a specific cancer type is associated with improved outcomes following checkpoint inhibitor therapy and, following from that observation, that the TMB threshold for benefit is not absolute (49). Because both MSI status (via MSIsensor) and TMB can be readily determined from targeted sequencing, correlative analysis of both MSI status and TMB in sarcoma immunotherapy trials on a subtype-specific basis is needed to inform our understanding.

Determining the clinical relevance of the landscape of genetic alterations in sarcomas described herein requires a further phase of investigation. Toward improved designs of clinical trials in sarcoma, which have often grouped multiple subtypes together despite significant inter- and intra-subtype genetic variability, future studies should investigate which genetic alterations result in functional effects. This may be particularly important in the subtypes we identified as having high entropy in their genomic clustering. That knowledge will enable the establishment of subtype *and* genotype-based trials to study the effect of novel agents in better defined biologic groups of tumors. The data we present herein and via an accompanying interactive database (cBioPortal link to be provided) will serve as a resource for the field to explore and compare subtype-specific alterations to facilitate this transition in approach.

## Methods

### Patient Cohort

This study was approved by the Institutional Review Board at Memorial Sloan Kettering Cancer Center (MSK). We identified patients with a diagnosis of soft tissue or bone sarcoma who had tumor and matched normal (white blood cell) tissue sequenced using the MSK-IMPACT assay through December 19, 2019 (5, 18). Tumors were sequenced using one of 3 versions of MSK-IMPACT, including 341, 410, or 468 genes, with results reported in the medical record. In patients with multiple samples, only one sample was included in the cohort; those collected earliest and of highest purity and highest average coverage were selected in that order of priority. Clinical characteristics such as patient age, sex, race, and metastatic versus primary site, were annotated per the standard MSK-IMPACT workflow (18).

### Histologic Analysis

Histologic diagnosis was annotated according to the standard MSK-IMPACT workflow. In the case of sarcomas characterized by canonical fusion events, the medical record was queried to ensure that the appropriate fusion event was detected and if not, the sample was reviewed with the assistance of an expert sarcoma pathologist. Similarly, samples harboring a canonical fusion but with a discordant pathologic diagnosis were further reviewed to assign the most appropriate diagnosis. Fusions other than those identified by MSK-IMPACT were annotated at the patient (not sample) level. Samples originally annotated as sarcoma or round cell sarcoma not otherwise specified, rhabdomyosarcoma (without further classification), spindle cell rhabdomyosarcoma, and fibrosarcoma underwent additional medical record review and, in some cases, pathology review to render the most accurate diagnosis possible. In some additional cases with ambiguity in subtype assignment, the diagnosis was updated upon further review by an expert pathologist. We further standardized diagnoses by mapping each tumor to a unique code from the OncoTree ontology (50) except for round cell sarcoma other (RCS (other)) and extraskeletal osteosarcoma, which were categories created for this study. Samples that could not be assigned to one of the Oncotree codes (n = 243) were excluded from our analysis cohort.

### Computational Genomic Analysis

Genomic alterations were annotated using the OncoKB precision oncology knowledge base, which identifies functionally relevant cancer variants and their potential clinical actionability (36). Except where otherwise specified in the text, variants of unknown significance, i.e. not labeled as *oncogenic*, *likely oncogenic*, or *predicted oncogenic* in OncoKB were excluded from the analysis. Therapeutically targetable somatic alterations were labeled using levels of clinical actionability defined in OncoKB, which range from level 1, FDA-recognized biomarkers of response to FDA-approved drugs, to level 4, biomarkers of hypothetical relevance based on compelling preclinical biological evidence. Analyses of alterations in oncogenic signaling pathways were performed using the set of pathway definitions previously curated by our group, which we expanded to include the DDR and epigenetic modifier pathways using additional templates curated from literature subtypes (24,51–53).

Tumor mutation burden (TMB) was computed as the total number of nonsynonymous mutations divided by the total number of base pairs sequenced per sample. The fraction of the genome altered (FGA) was defined as the fraction of genome with log_2_ copy number gain > 0.2 or loss < −0.2 relative to the size of the genome for which copy number was profiled. We computed MSIsensor scores for all samples in the cohort and used a threshold of MSIsensor score ≥ 10 to identify tumors with microsatellite instability (MSI-high) (54). MSI-high was confirmed by a PCR-based assay (Idylla). MSIsensor ≥ 3 and < 10 were labeled indeterminate and samples that did not meet quality control for assigning MSI status were labeled do not report (DNR).

Allele-specific copy number estimates at both the gene and chromosome arm levels were computed using the FACETS (Fraction and Allele-Specific Copy Number Estimates from Tumor Sequencing) algorithm, which also provided purity-corrected segmentation files and allowed identification of whole-genome duplication events (55). FACETS output was also used to infer the cancer cell fraction associated with individual mutations for clonality analyses. Significantly recurrently mutated genes were identified using the MuSic and MutSigCV 1.4 algorithms, with a threshold q-value of 0.1 (56, 57). Dimensionality reduction was performed using Uniform Manifold Approximation and Projection (UMAP) (http://arxiv.org/abs/1802.03426) and clusters were identified using Hierarchical Density-Based Spatial Clustering of Applications with Noise (HDBSCAN) (58). Shannon entropy was calculated from observed cluster assignment by subtype and reported in natural units.

Mutational signatures for samples with ≥ 15 synonymous and nonsynonymous single nucleotide variants (SNVs) were extracted using the COSMIC v3 catalog of exome reference signatures and default parameters (59) (https://github.com/mskcc/tempoSig). For mutational signatures to be considered detectable, we required a p-value < 0.05 and a minimum of 1 observed mutation attributed to the signature, where the number of observed mutations was defined as the observed mutational signature fraction multiplied by the number of SNVs per sample.

### Data Availability

All clinical and genomic data described in this manuscript will be accessible online and publicly available for bulk download through the cBioPortal for Cancer Genomics (60).

## Data Availability

All clinical and genomic data described in this manuscript will be accessible online and publicly available for bulk download through the cBioPortal for Cancer Genomics.

## Author contributions

Conception and design: Nacev, Sanchez-Vega, Gounder, Bowler, Singer, Tap

Development of methodology: Nacev, Sanchez-Vega, Smith, Shi, Tang, Socci

Data collection: Nacev, Antonescu, Zehir, Gounder, Chi, D’Angelo, Dickson, Kelly, Keohan, Movva, Thornton, Meyers, Wexler, Slotkin, Glade Bender, Shukla, Hensley, Healey, La Quaglia, Crago, Yoon, Untch, Chiang, Agaram, Hameed, Berger, Solit, Ladanyi, Singer, Tap

Data curation: Nacev, Sanchez-Vega, Smith, Antonescu, Bowler

Resources: Sanchez-Vega, Smith, Shi, Tang, Zehir, Berger, Schultz

Analysis and interpretation of data: Nacev, Sanchez-Vega, Smith, Antonescu, Rosenbaum, Shi, Tang, Socci, Rana, Gularte-Mérida, Zehir, Luthra, Jadeja, Okada, Strong, Stoller, Berger, Solit, Schultz, Singer, Tap

Drafting the manuscript – Nacev, Sanchez-Vega, Rosenbaum, Singer, Tap

Review and revision of the manuscript: All authors

Visualization of data: Sanchez-Vega, Smith, Shi, Tang

Study supervision and funding acquisition: Singer, Tap

## Supplementary Tables

Supplementary Table 1. Master sample dataset. MSI, microsatellite instability; FGA, fraction of the genome altered; OS, overall survival; FACETS, Fraction and Allele-Specific Copy Number Estimates from Tumor Sequencing algorithm; QC, quality control; CNA, copy number alterations; WGD, whole genome doubling.

Supplementary Table 2. Patient characteristics by subtype.

Supplementary Table 3. Number of oncogenic fusion events in individual genes in each subtype.

Supplementary Table 4. Pathway gene composition. CC, cell cycle; EPI, epigenetic.

Supplementary Table 5. Biochemical function/complex assignments for epigenetic pathway genes.

**Supplementary Figure 1.**
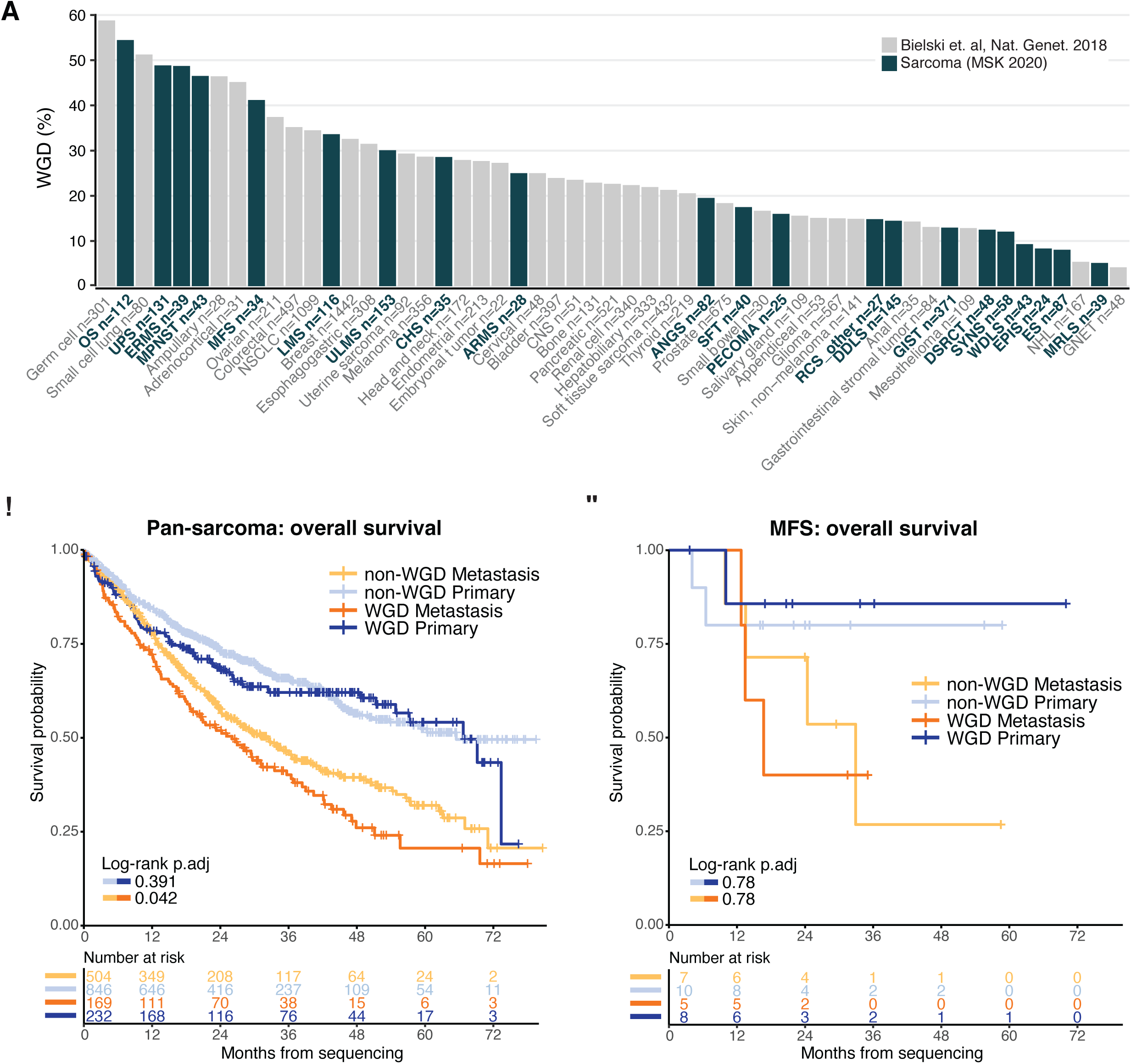
Whole genome doubling (WGD) and survival probability. **A**, Frequency of WGD by subtype (green) compared to other cancers (all available samples) (gray). **B-C**, Overall survival based on WGD status within metastatic and primary tumor cohorts in **B**, all sarcoma subtypes; **C**, MFS.

**Supplementary Figure 2.**
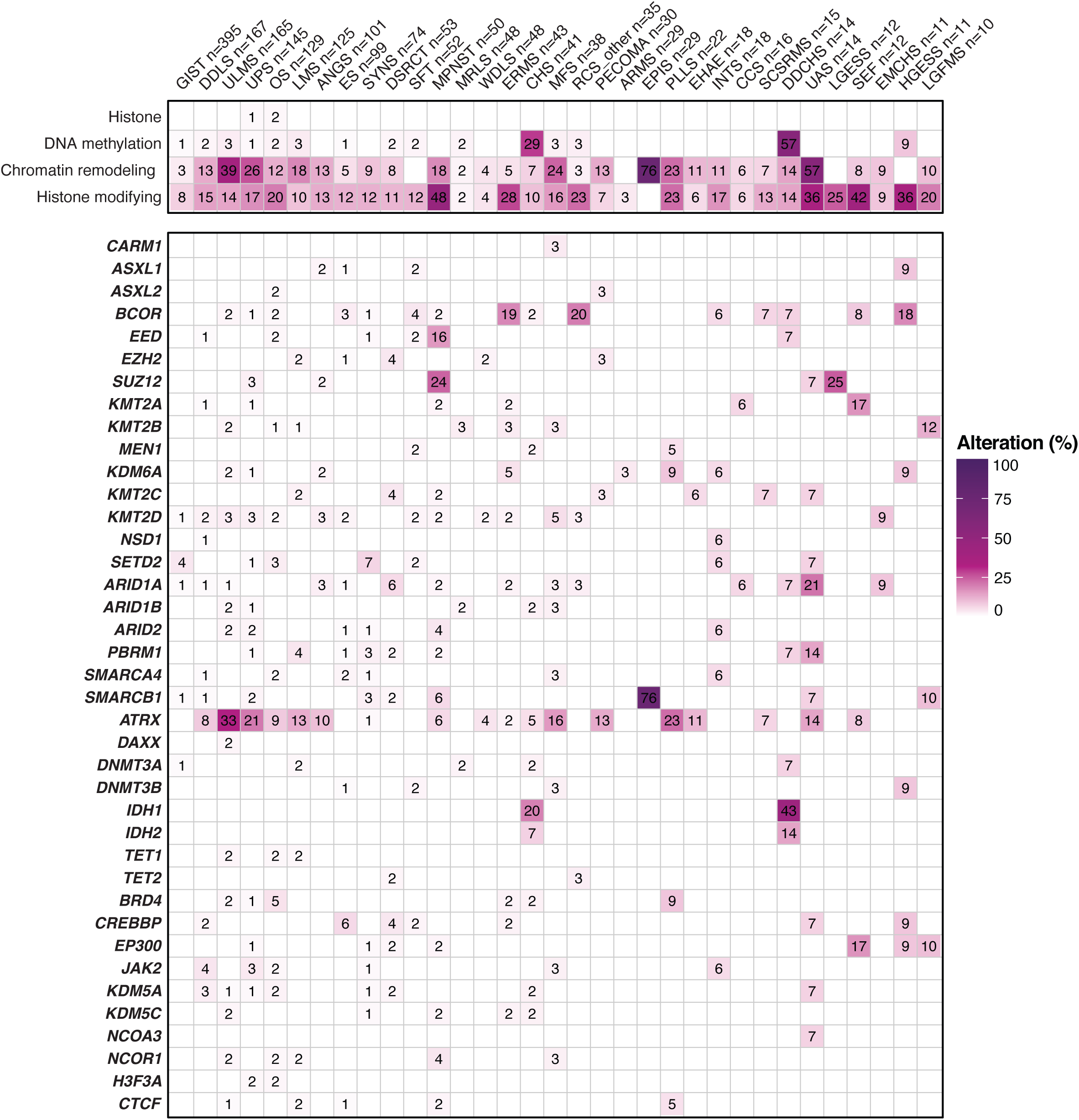
Oncogenic epigenetic pathway alterations. Frequency of oncogenic alterations in specific epigenetic pathway genes in each subtype with ≥ 10 samples. Top box, aggregate number of alterations in each gene family/biochemical process.

**Supplementary Figure 3.**
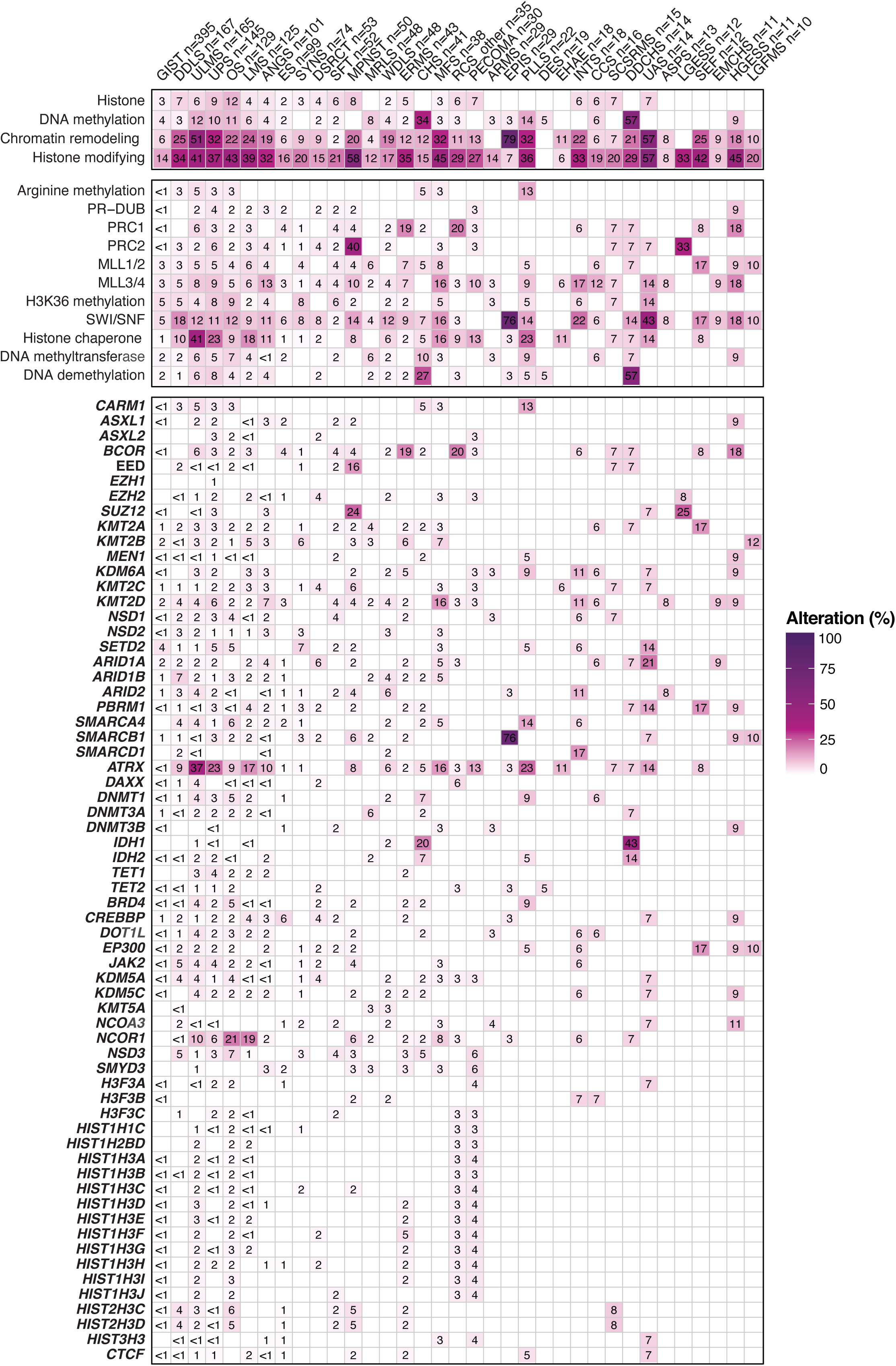
All epigenetic pathway alterations, including variants of unknown significance (VUS). Frequency of somatic alterations in epigenetic pathway genes in each subtype with ≥ 10 samples. Top boxes, aggregate number of alterations in each gene family and biochemical process.

