## Supplemental Table 1 for "Clinical sequencing of soft tissue and bone sarcomas delineates diverse genomic landscapes and potential therapeutic targets"

| | SAMPLES | AGE | | SEX | | SAMPLE TYPE | |
|---|---|---|---|---|---|---|---|
| | **Count** | **Mean ± SD** | **Range** | **Male** | **Female** | **Metastatic** | **Primary** |
| **TOTAL** | 2138 | 49.3 ± 20.0 | 0–90 | 1040 | 1098 | 790 | 1348 |
| **GIST** | 395 | 58.5 ± 14.2 | 11–90 | 213 | 182 | 123 | 272 |
| **DDLS** | 167 | 62.1 ± 11.7 | 29–90 | 115 | 52 | 58 | 109 |
| **ULMS** | 165 | 57.2 ± 10.3 | 30–83 | 0 | 165 | 91 | 74 |
| **UPS** | 145 | 55.6 ± 18.1 | 3–87 | 74 | 71 | 52 | 93 |
| **OS** | 129 | 26.1 ± 16.6 | 8–78 | 79 | 50 | 52 | 77 |
| **LMS** | 125 | 57.8 ± 14.6 | 7–83 | 49 | 76 | 71 | 54 |
| **ANGS** | 101 | 58.6 ± 16.7 | 15–89 | 41 | 60 | 24 | 77 |
| **ES** | 99 | 26.9 ± 16.1 | 2–79 | 60 | 39 | 28 | 71 |
| **SYNS** | 74 | 40.7 ± 16.0 | 7–85 | 36 | 38 | 24 | 50 |
| **DSRCT** | 53 | 23.4 ± 9.4 | 8–48 | 48 | 5 | 24 | 29 |
| **SFT** | 52 | 60.2 ± 11.0 | 39–80 | 25 | 27 | 22 | 30 |
| **MPNST** | 50 | 43.7 ± 20.2 | 11–89 | 30 | 20 | 12 | 38 |
| **MRLS** | 48 | 44.7 ± 13.4 | 13–67 | 29 | 19 | 16 | 32 |
| **WDLS** | 48 | 57.8 ± 9.9 | 34–74 | 27 | 21 | 25 | 23 |
| **ERMS** | 43 | 15.5 ± 16.8 | 1–70 | 20 | 23 | 7 | 36 |
| **CHS** | 41 | 47.9 ± 15.4 | 16–76 | 26 | 15 | 16 | 25 |
| **MFS** | 38 | 64.5 ± 12.6 | 31–80 | 21 | 17 | 13 | 25 |
| **RCS_other** | 35 | 27.8 ± 18.0 | 0–70 | 14 | 21 | 3 | 32 |
| **PECOMA** | 30 | 55.9 ± 15.0 | 14–76 | 7 | 23 | 15 | 15 |
| **ARMS** | 29 | 23.5 ± 16.4 | 1–67 | 16 | 13 | 13 | 16 |
| **EPIS** | 29 | 38.5 ± 15.2 | 16–67 | 14 | 15 | 9 | 20 |
| **PLLS** | 22 | 53.0 ± 18.9 | 12–87 | 10 | 12 | 8 | 14 |
